# Multimodal Imaging Identifies Cardiac Remodeling Phenotype With Reduced Exercise Capacity in Repaired Tetralogy of Fallot

**DOI:** 10.64898/2026.08.23.26361146

**Authors:** Bryan P Mosher, Jeffrey W Christle, Jason V Tso, Euan A Ashley, Daniel E Clark

**Affiliations:** Stanford Cardiovascular Institute, Stanford, CA. USA; Department of Medicine, Stanford, CA. USA; Division of Cardiovascular Medicine, Stanford, CA. USA; Adult Congenital Heart Disease, Stanford, CA. USA

**Keywords:** cardiac magnetic resonance, cardiopulmonary exercise testing, diastolic dysfunction, echocardiography, left atrial volume index, right ventricular function

## Abstract

**Background:** Exercise intolerance is common in adults with repaired tetralogy of Fallot (rTOF) despite preserved left ventricular ejection fraction (LVEF ≥50%). Whether reduced exercise capacity is associated with early cardiac remodeling remains unclear.

**Objectives:** To determine whether reduced exercise capacity in rTOF with preserved LVEF is associated with diastolic dysfunction, atrial remodeling, right ventricular (RV) dysfunction, and myocardial fibrosis.

**Methods:** We retrospectively studied adults with rTOF and preserved LVEF who underwent cardiopulmonary exercise testing (CPET) and transthoracic echocardiography (TTE) and/or cardiac MRI (CMR) within 18 months. Exercise capacity was assessed by percent-predicted peak VO2 (ppVO2). Diastolic function and atrial remodeling were evaluated by TTE, and CMR assessed RV function and myocardial fibrosis.

**Results:** Reduced exercise capacity was associated with larger left atrial volume index (LAVI; p < 0.001), elevated E/e′ and reduced e′ velocity (both p < 0.05), and reduced RV systolic function (p < 0.001). LAVI correlated inversely with ppVO2 (ρ = −0.27, p = 0.003). A composite diastolic dysfunction score showed a graded relationship with exercise capacity, with patients exhibiting ≥2 abnormalities having lower ppVO2 than those with ≤1 abnormality (both p < 0.01). In contrast, pulmonary regurgitation (PR) severity and myocardial fibrosis by late gadolinium enhancement (LGE) were not associated with exercise capacity.

**Conclusions:** In adults with rTOF and preserved LVEF, reduced exercise capacity is associated with atrial remodeling, diastolic dysfunction, and RV dysfunction despite the absence of overt myocardial fibrosis. This suggests that multimodal imaging identifies an imaging-defined cardiac remodeling phenotype associated with early functional impairment.

## Introduction

Tetralogy of Fallot (TOF) is the most common cyanotic congenital heart disease (CHD), accounting for 7-10% of all congenital heart defects and affecting approximately 1 in 3,500 births.^1^ Significant advancements in the diagnosis and management of TOF have resulted in improved survival and a rapidly growing population of adults with repaired TOF (rTOF).^2^ Despite these advances, rTOF patients face significant lifetime late morbidity and mortality.^2^ Diminished exercise capacity is a prevalent and potentially debilitating morbidity in patients with rTOF.^2^ Low peak oxygen uptake (VO2) on cardiopulmonary exercise testing (CPET) less than 36% predicted is correlated with mortality in adults with rTOF.^1^ There is a need to increase our understanding of predictors of low VO2 and in this population to allow for early intervention.

Beginning in early adulthood, patients with rTOF often experience exercise intolerance and reduction in exercise capacity despite preserved left ventricular systolic function (LVEF; ≥50%).^3^ This dissociation suggests that early impairment in functional reserve may precede LV dysfunction and structural myocardial disease. However, the mechanisms underlying this early decline remain incompletely understood. In many patients, traditional markers of myocardial disease, including systolic dysfunction and myocardial fibrosis, are absent at the time when functional limitation first becomes apparent.^2^ This raises the possibility that an intermediate remodeling phenotype exists prior to the development of irreversible structural injury.

In acquired cardiovascular disease, particularly heart failure with preserved ejection fraction (HFpEF), early myocardial remodeling is characterized by impaired ventricular relaxation, elevated filling pressures, and left atrial remodeling, often occurring before overt systolic dysfunction or fibrosis.^4^ Adults with CHD also develop HFpEF^5^ and may represent a unique human model for studying these early transitions, as they are exposed to decades of chronic hemodynamic stress from abnormal loading conditions.

Repaired TOF is an especially relevant population in which to investigate this phenomenon. Chronic RV volume loading and altered ventricular interaction impose sustained physiologic stress, yet many patients maintain preserved LV systolic function for years. This creates an opportunity to examine early myocardial adaptations that may precede irreversible structural disease.

Accordingly, this study examines whether reduced exercise capacity in adults with rTOF and preserved LVEF is associated with early markers of myocardial remodeling. Specifically, we evaluate the relationship between CPET-derived percent-predicted peak VO2 (ppVO2), a measure of functional reserve, and transthoracic echocardiographic (TTE) indices of diastolic function, alongside cardiac magnetic resonance (CMR)-derived measures of ventricular function and myocardial fibrosis, to determine whether functional limitation precedes overt structural disease.

We hypothesized that among adults with rTOF and preserved LVEF, reduced ppVO2 reflects an early myocardial remodeling phenotype characterized by impaired diastolic function, elevated filling pressures, and left atrial enlargement, occurring prior to the development of overt myocardial fibrosis.

## Methods

### Study Population

We conducted a retrospective cohort study of 144 adults with rTOF and preserved LVEF followed at a tertiary adult CHD center (Table 1). Patients were included if they underwent CPET and TTE within 18 months of each other. The median interval between CPET and TTE was 177 days (IQR 0-364 days). CPET studies with inadequate effort, defined as a respiratory exchange ratio <1.05, were excluded. For the primary analysis, patients were restricted to those with preserved LVEF in order to isolate early myocardial remodeling prior to the development of overt ventricular dysfunction. No exclusions were made on the basis of clinical outcomes. When available, CMR imaging performed within 18 months of CPET was included for assessment of ventricular function, tricuspid regurgitation (TR), pulmonary regurgitation (PR), and myocardial fibrosis. The institutional review board approved this study (IRB 78353).

**Table 1.** Baseline Characteristics.

| **Variable** | **Overall** |
| --- | --- |
| Age (years) | 39.0 (31.0–50.0) |
| Male sex, n (%) | 77 (54%) |
| Female sex, n (%) | 67 (46%) |
| BMI (kg/m<sup>2</sup>) | 24.2 (22.1–27.5) |
| Peak VO<sub>2</sub> (mL/kg/min) | 25.5 (21.0–32.7) |
| Peak VO<sub>2</sub> (% predicted) | 69.0 (58.0–81.0) |
| RVEF category | 0: 99 (68%); 1: 32 (22%); 2: 13 (9%); 3: 1 (1%) |
| LAVI (mL/m<sup>2</sup>) | 21.6 (18.5–26.8) |
| E/e' | 10.3 (7.8–12.7) |
| Lateral e' (cm/s) | 11.9 (9.9–14.6) |
| TR V<sub>max</sub> (m/s) | 2.7 (2.5–3.0) |
| Diastolic dysfunction score distribution | 0: 94 (58%); 1: 50 (31%); 2: 14 (9%); 3: 4 (2%) |
| LGE on CMR | 19/107 (18%) |

Values are median (IQR) or n (%). RVEF categories are defined as 0 = normal, 1 = mild, 2 = moderate, and 3 = severe dysfunction. The diastolic dysfunction score (0–4) is the sum of LAVI >34 mL/m<sup>2</sup>, E/e' >14, lateral e' <10 cm/s, and TR V<sub>max</sub> >2.8 m/s. Abbreviations: BMI = body mass index; CMR = cardiac magnetic resonance; E/e' = ratio of early transmitral flow velocity to early diastolic mitral annular velocity; IQR = interquartile range; LAVI = left atrial volume index; LGE = late gadolinium enhancement; RVEF = right ventricular ejection fraction; TR V<sub>max</sub> = peak tricuspid regurgitation velocity; VO<sub>2</sub> = oxygen consumption.

### Functional Assessment

Exercise capacity was assessed using peak oxygen uptake (VO2) obtained during CPET. The primary outcome was ppVO2, calculated according to the Fitness Registry and the Importance of Exercise National Database (FRIEND).^6^ For patients with multiple CPETs, the most recent CPET meeting the study inclusion criterion of a TTE within 18 months was used for analysis. When available, CMR studies performed within 18 months of CPET and TTE were included for secondary analyses.

### Echocardiographic Assessment

Echocardiographic measurements were obtained from clinically obtained TTE studies according to institutional standard, Intersocietal Accreditation Commission protocols and included indices of diastolic function and cardiac remodeling, and ventricular function of LV (quantitative) and RV (qualitative). Diastolic function was assessed using mitral inflow parameters, including E/A ratio, tissue Doppler imaging of the mitral annulus (lateral and septal e′ velocities), and the ratio of early mitral inflow velocity to annular velocity (E/e′), calculated using average annular velocities. Left atrial remodeling was assessed using left atrial volume index (LAVI). Right-sided parameters included peak tricuspid regurgitation velocity (TR V_max_) and qualitative assessment of TR severity.

Given the known limitations of septal annular velocities in repaired tetralogy of Fallot, including the effects of ventricular interaction, conduction delay, and prior surgical intervention, lateral e′ was prioritized in primary analyses, with septal e′ used in secondary analyses.

### Cardiac Magnetic Resonance

When available, CMR data were used to assess left and right ventricular systolic function and the presence of myocardial fibrosis using late gadolinium enhancement (LGE). TR and PR were quantified using phase-contrast imaging when available and reported as regurgitant fraction.

### Definition of Diastolic Dysfunction

Diastolic dysfunction was assessed using both individual TTE parameters and a composite diastolic dysfunction score. The composite score incorporated established guideline-recommended parameters, including LAVI >34 mL/m², average E/e′ >14, lateral e′ <10 cm/s, and TR V_max_ >2.8 m/s.^7^ Each abnormal parameter contributed one point, yielding a total score ranging from 0 to 4. For categorical analyses, individual diastolic parameters were dichotomized using these clinically established thresholds.

### Statistical Analysis

Given the non-normal distribution of physiologic variables, non-parametric methods were used for primary analyses. For further analyses, continuous variables are presented as mean ± standard deviation or median with interquartile range, as appropriate, and categorical variables are presented as counts and percentages. Group comparisons were performed using the Wilcoxon rank-sum test, and associations between continuous variables were assessed using Spearman correlation coefficients. The primary outcome was ppVO2. Analyses focused on the relationship between exercise capacity and markers of diastolic function, atrial remodeling, ventricular function, and myocardial fibrosis. A two-sided p < 0.05 was considered statistically significant. A multivariable linear regression model including LAVI and CMR-derived RVEF was constructed to determine whether atrial remodeling and RV systolic function were independently associated with ppVO2.

## Results

In the total population, median ppVO2 was 69.0% (IQR 58.0–81.0), and median absolute peak VO2 was 25.5 mL/kg/min (IQR 21.0–32.7), indicating substantial heterogeneity in exercise capacity despite preserved LVEF. The majority of patients (107; 74%) had a CMR within 18 months of CPET.

### Associations Between Diastolic Function and Exercise Capacity

Markers of diastolic dysfunction were significantly associated with reduced exercise capacity. Patients with enlarged LAVI (>34 mL/m²) had significantly lower ppVO2 compared with those with normal LAVI (52.9 ± 13.3% vs 71.2 ± 15.4%, p = 0.0007; Figure 1A). LAVI also demonstrated a modest inverse correlation with ppVO2 (Spearman ρ = −0.267, p = 0.0025; n = 126). Similarly, elevated E/e′ was associated with reduced exercise capacity, with patients demonstrating E/e′ >14 having lower ppVO2 compared with those with normal filling pressures (60.8 ± 16.6 vs 73.2 ± 15.2, p < 0.05) (Figure 1B). In parallel, reduced myocardial relaxation velocity (e′ <10 cm/s) was associated with impaired exercise capacity (66.0 ± 18.1 ppVO2 vs 73.9 ± 14.9 ppVO2, p < 0.05) (Figure 1C). In contrast, mitral inflow E/A ratio was not significantly associated with ppVO2, suggesting that conventional Doppler indices alone may be less sensitive markers of early functional limitation. TR Vmax demonstrated a weak inverse correlation with ppVO2 (Spearman ρ = −0.19, p = 0.034, n = 120) (Figure 1D).

**Figure 1.**
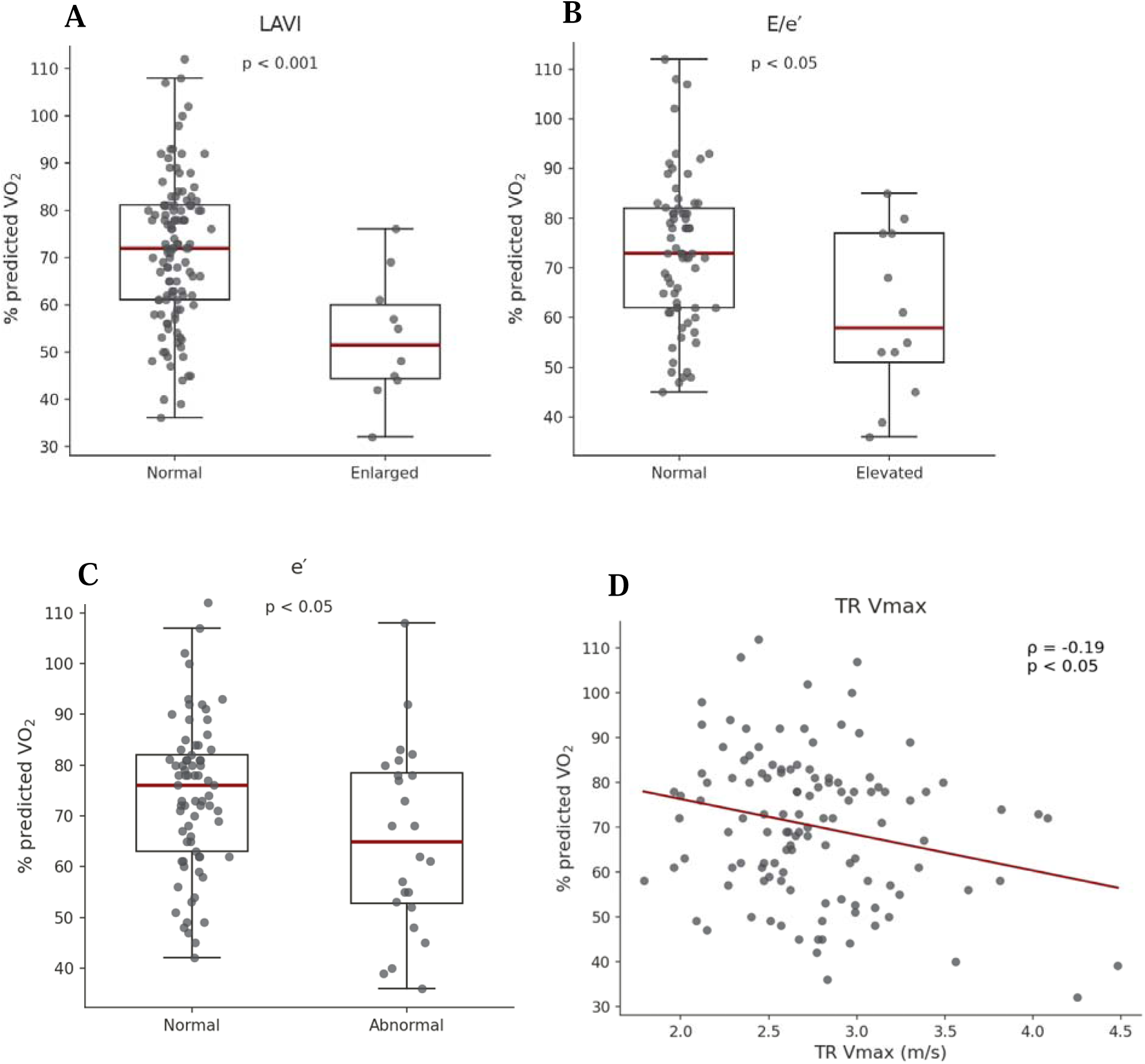
Echocardiographic Markers and Exercise Capacity. **(A)** Patients with enlarged left atrial volume index (LAVI >34 mL/m²) demonstrated lower ppVO2 than those with normal LAVI (p < 0.001). **(B)** Elevated E/e′ (>14) was associated with lower ppVO2 than normal E/e′ (≤14) (p < 0.05). **(C)** Reduced lateral mitral annular early diastolic velocity (e′ <10 cm/s) was associated with lower ppVO2 than normal e′ (≥10 cm/s) (p < 0.05). **(D)** Peak tricuspid regurgitation velocity (TR Vmax) demonstrated a weak inverse correlation with ppVO2 (Spearman ρ = −0.19; p < 0.05). Box plots show the median and interquartile range; individual points represent individual patients. Abbreviations: LAVI = left atrial volume index; LVEF = left ventricular ejection fraction; ppVO2 = percent-predicted peak oxygen consumption; TR Vmax = peak tricuspid regurgitation velocity.

### Composite Diastolic Dysfunction and Functional Capacity

A composite diastolic dysfunction score incorporating LAVI, E/e′, e′, and TR Vmax demonstrated a graded relationship with exercise capacity. Patients with ≥2 abnormal parameters had significantly lower ppVO2 than those with 0 or 1 abnormality (Figure 2A). The most common abnormality pattern was TR Vmax >2.8 m/s without another parameter classified as abnormal (n = 31), followed by e′ <10 cm/s without another parameter classified as abnormal (n = 10). Additional patients demonstrated multiple combinations of abnormal diastolic parameters, although no single combination predominated (Figure 2B).

**Figure 2.**
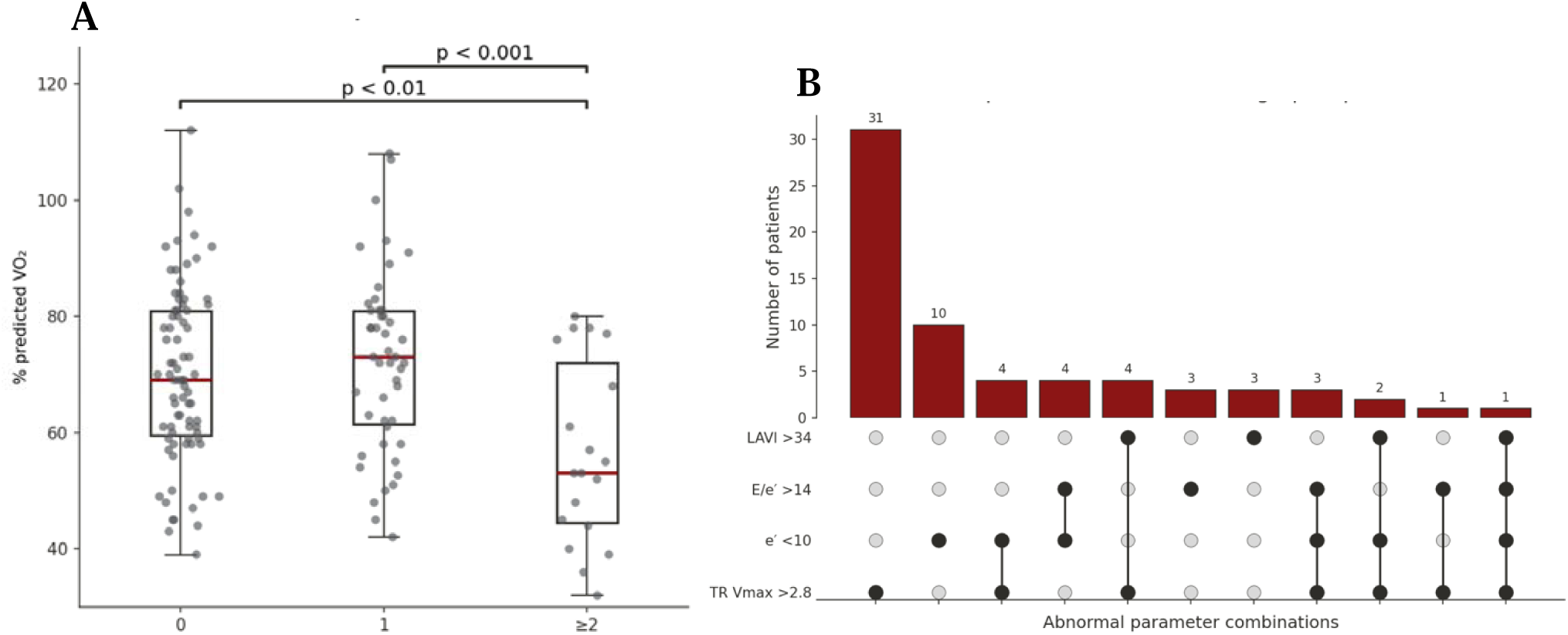
Diastolic Remodeling Burden and Exercise Capacity. **(A)** Patients with a greater burden of abnormal echocardiographic diastolic parameters demonstrated progressively lower ppVO2. Individuals with ≥2 abnormalities had significantly lower exercise capacity than those with 0 or 1 abnormality. **(B)** UpSet plot showing the frequency and co-occurrence of abnormal LAVI, E/e′, lateral e′, and TR Vmax. Multiple combinations of abnormalities were observed, supporting the presence of a composite remodeling phenotype. Abbreviations: LAVI = left atrial volume index; ppVO2 = percent-predicted peak oxygen consumption; TR Vmax = peak tricuspid regurgitation velocity.

### Pulmonary Valve Regurgitation and Exercise Capacity

PR fraction measured by CMR was not associated with ppVO2 (ρ = -0.08, p = 0.414; Figure 4A). Similarly, CMR-derived RV end-diastolic volume index was not associated with ppVO2 (ρ = −0.04, p = 0.711; n = 106). In contrast, worsening RV systolic dysfunction was associated with lower ppVO2 (p < 0.001; Figure 3).

**Figure 3.**
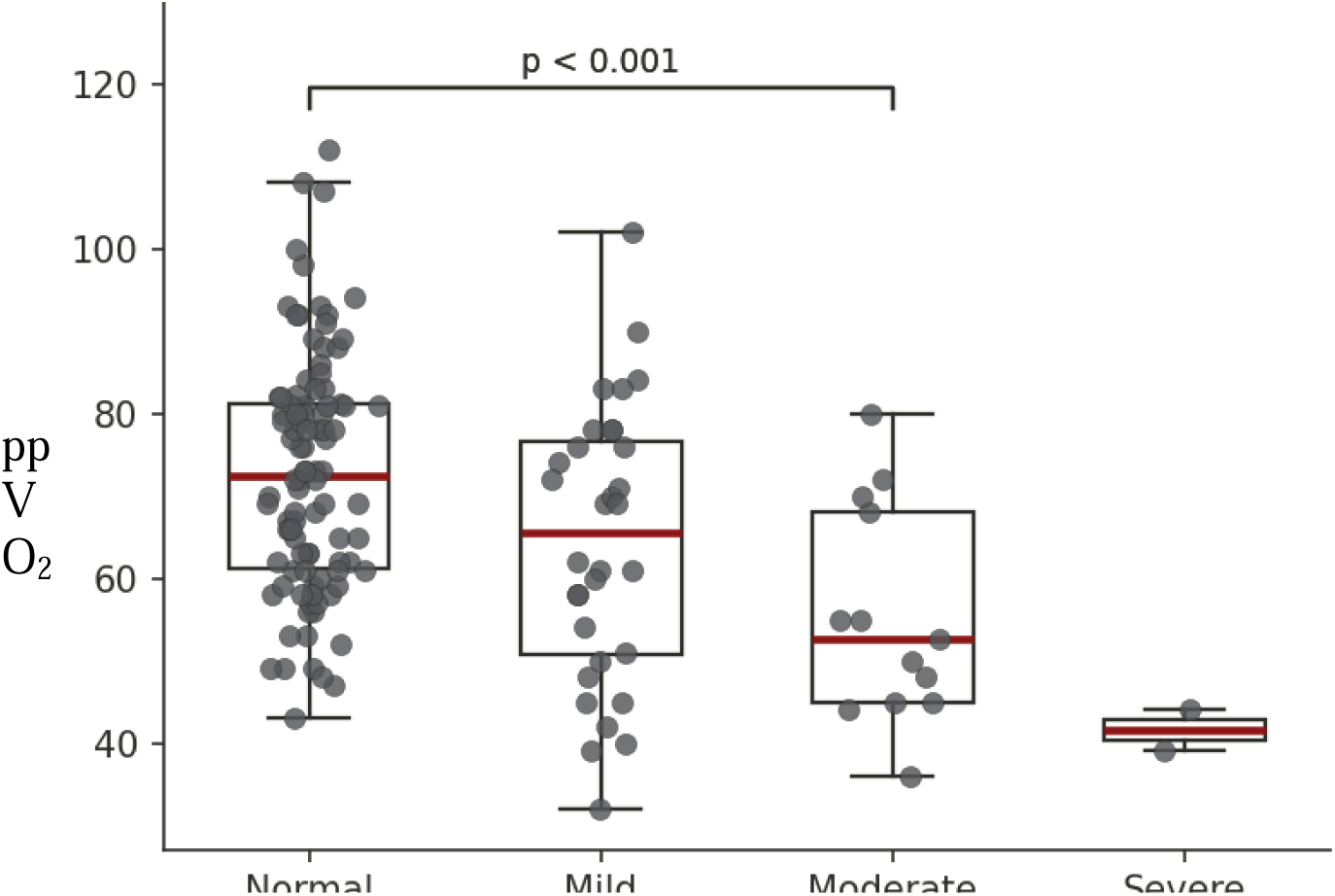
Right Ventricular Systolic Dysfunction and Exercise Capacity. CMR-derived RV systolic dysfunction was associated with reduced exercise capacity despite preserved LVEF. Patients with worsening RV systolic dysfunction demonstrated progressively lower ppVO2. Individuals with moderate RV dysfunction had significantly lower exercise capacity than those with normal RV function (p < 0.001). Box plots show the median and interquartile range; individual points represent individual patients. Abbreviations: CMR = cardiac magnetic resonance; LVEF = left ventricular ejection fraction; ppVO2 = percent-predicted peak oxygen consumption; RV = right ventricle/right ventricular.

**Figure 4.**
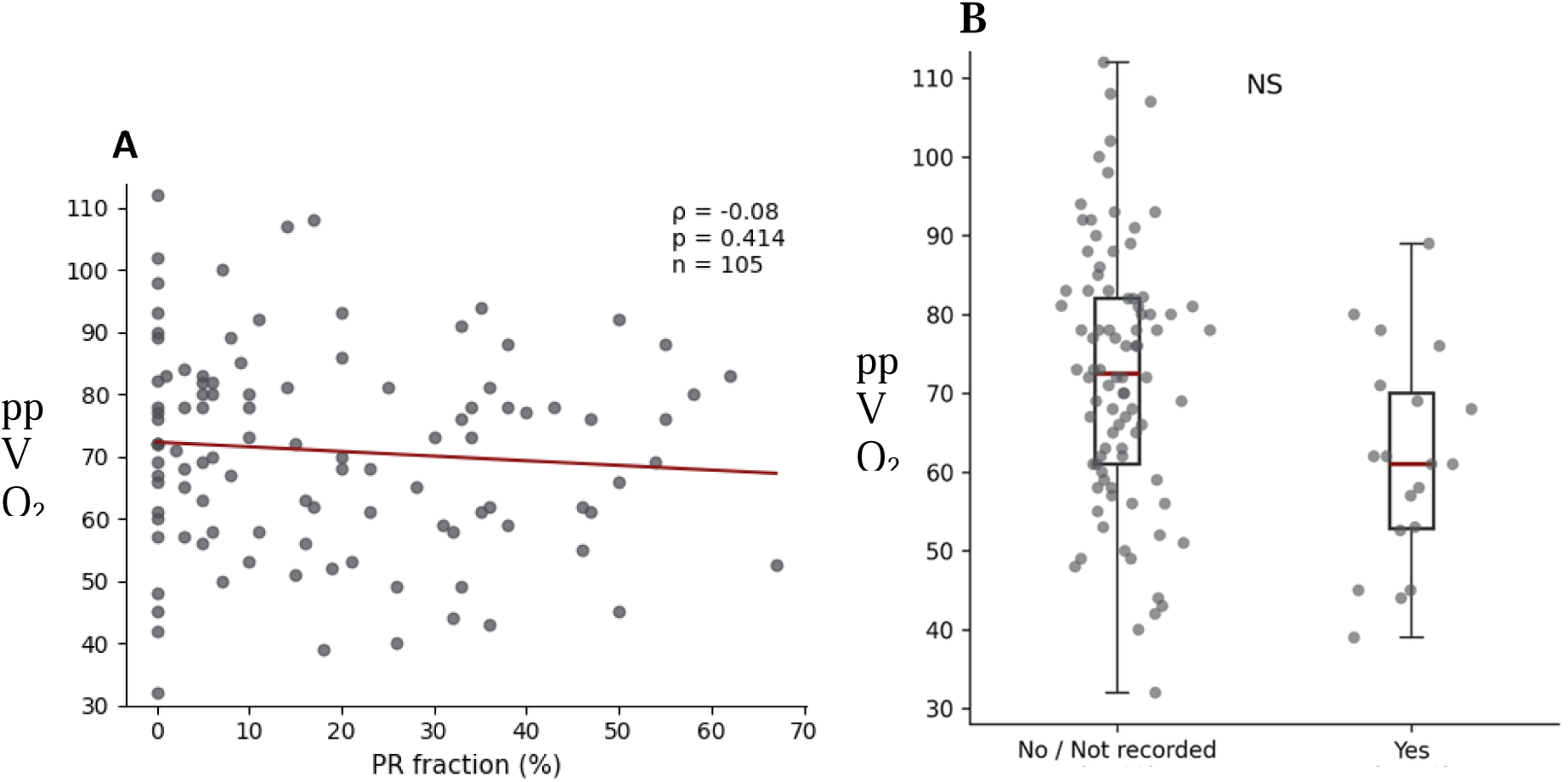
Pulmonary Regurgitation, Myocardial Fibrosis, and Exercise Capacity. **(A)** PR fraction was not associated with ppVO2 (Spearman ρ = −0.08; p = 0.414). **(B)** Among 107 patients with available CMR data, the presence of LGE was not associated with ppVO2. Box plots show the median and interquartile range; individual points represent individual patients. Abbreviations: CMR = cardiac magnetic resonance; LGE = late gadolinium enhancement; NS = not significant; ppVO2 = percent-predicted peak oxygen consumption; PR = pulmonary regurgitation.

In a multivariable linear regression including both LAVI and CMR-derived RVEF, larger LAVI (β = −0.65, 95% CI −1.12 to −0.19; p = 0.007) and higher RVEF (β = 0.62, 95% CI 0.19 to 1.06; p = 0.006) were independently associated with ppVO2 (R² = 0.17).

### Fibrosis by LGE on CMR and Exercise Capacity

Among 107 patients with available CMR data, 19 (18%) demonstrated LGE. The presence of LGE was not associated with ppVO2 (Figure 4B).

## Discussion

In this cohort of adults with rTOF and preserved LVEF, reduced exercise capacity was associated with markers of diastolic dysfunction and atrial remodeling. Left atrial enlargement (LAVI), elevated filling pressures (E/e′), and impaired myocardial relaxation (reduced e′ velocity) were each associated with lower ppVO2. A composite diastolic dysfunction score demonstrated a graded relationship with exercise capacity, supporting an additive effect of multiple abnormalities on exercise capacity. In contrast, PR severity was not associated with exercise capacity, whereas worsening RV systolic dysfunction was associated with lower ppVO2. A minority of patients demonstrated fibrosis on CMR. Collectively, these findings support the presence of an early myocardial remodeling phenotype in rTOF characterized by diastolic dysfunction and atrial remodeling despite preserved systolic function. The proposed imaging-defined cardiac remodeling phenotype and its association with reduced exercise capacity are summarized in the Central Illustration.

Exercise intolerance is common in rTOF, yet its mechanistic basis remains incompletely understood. Prior studies have emphasized RV volume overload and systolic dysfunction as primary drivers of reduced exercise capacity.^1,8,9^ However, our findings suggest that abnormalities in diastolic function may also play a central role in early functional limitation. The associations observed between impaired relaxation (reduced e′), elevated filling pressures (E/e′), and reduced ppVO2 are consistent with mechanisms described in HFpEF, where impaired ventricular filling, reduced compliance, and elevated filling pressures limit stroke volume augmentation during exercise.^4,10,11^ These parallels suggest that rTOF may share common physiologic pathways with HFpEF, despite fundamentally different underlying etiologies. Notably, conventional Doppler indices such as the E/A ratio were not associated with exercise capacity, underscoring the limitations of single-parameter assessments and reinforcing the importance of integrative evaluation of diastolic function.

An additional mechanism that may contribute to these findings is ventriculo-ventricular interaction. In rTOF, chronic RV volume loading and altered RV geometry can impair LV filling through septal displacement and pericardial constraint.^12,13^ These interactions may contribute to abnormalities in diastolic filling and exercise capacity despite preserved LVEF. Although ventricular interaction was not directly assessed in this study, the coexistence of RV dysfunction, diastolic abnormalities, and reduced exercise capacity is consistent with this physiologic framework.

Importantly, LAVI and CMR-derived RVEF remained independently associated with exercise capacity in a multivariable model. This finding suggests that atrial remodeling and RV systolic dysfunction provide complementary information regarding early myocardial remodeling rather than reflecting the same underlying physiologic abnormality.

Left atrial volume index (LAVI) demonstrated one of the strongest associations with ppVO_2_ in this study. Left atrial enlargement reflects the cumulative burden of chronically elevated filling pressures and has been shown in multiple populations to serve as a robust marker of diastolic dysfunction and adverse cardiovascular outcomes.^14,15^ Figure 2B demonstrates that abnormalities in left atrial size, myocardial relaxation, filling pressure, and TR Vmax occurred in multiple combinations rather than following a single stereotyped pattern. In contrast, isolated abnormalities in the other diastolic parameters were uncommon. This pattern suggests that atrial remodeling may represent one of the earliest detectable manifestations of adverse myocardial adaptation in rTOF, preceding the development of more widespread abnormalities in ventricular relaxation, filling pressures, or pulmonary hemodynamics. The strong association between LAVI and exercise capacity further supports the potential role of atrial remodeling as a marker of functional impairment.

In this context, LAVI likely reflects cumulative hemodynamic exposure over time, whereas E/e′ provides a dynamic estimate of filling pressures at a single time point. These findings highlight left atrial enlargement as a particularly sensitive marker of early myocardial remodeling in rTOF and suggest that atrial metrics may be more informative than traditional ventricular measures in identifying subclinical disease.

The composite diastolic dysfunction score demonstrated a graded relationship with exercise capacity, with progressively lower ppVO2 observed as the burden of abnormal parameters increased (Figure 2A). Patients with at least two abnormalities had significantly lower exercise capacity than those with zero or one abnormality. Figure 2B further demonstrates that abnormalities in left atrial size, myocardial relaxation, filling pressure, and TR Vmax occurred in multiple combinations rather than as a single stereotyped echocardiographic pattern.

Although the continuous correlation between the composite score and ppVO2 was modest, the categorical relationship was robust, suggesting that the accumulated burden of abnormalities may be more clinically meaningful than any individual parameter. These findings support an integrative approach to identifying early cardiac remodeling in rTOF. This is consistent with prior work demonstrating that integrated diastolic indices outperform individual measures in capturing the complexity of myocardial physiology.^16,17^ Importantly, this approach aligns with guideline-recommended frameworks for the assessment of diastolic dysfunction, which emphasize the integration of multiple echocardiographic parameters rather than reliance on a single metric.^7^

A key implication of these findings is the dissociation between functional limitation and overt structural myocardial disease. Despite preserved LVEF and the absence of strong associations with myocardial fibrosis or PR, substantial variability in exercise capacity was observed. These findings support a model in which functional impairment is associated with early myocardial remodeling despite the absence of overt structural myocardial abnormalities detectable by imaging. In this context, rTOF may represent a model of early myocardial remodeling analogous to preclinical HFpEF, where abnormalities in relaxation and filling precede overt systolic dysfunction and fibrosis. These observations suggest that functional and physiologic assessments may be helpful in the detection of early disease.

Although RV systolic dysfunction was associated with reduced exercise capacity, PR severity was not. Similarly, CMR-derived RV end-diastolic volume index was not associated with ppVO2, suggesting that impaired RV systolic function may be more closely related to exercise limitation than the degree of RV dilation alone. These findings indicate that volume loading alone does not fully explain functional limitation and are consistent with prior studies demonstrating that exercise capacity in rTOF is multifactorial and not solely determined by RV volume overload.^2,18^ RV dysfunction may coexist with other physiologic abnormalities that contribute to reduced exercise capacity. These findings further support the concept that diastolic abnormalities and atrial remodeling may represent earlier and more sensitive markers of disease progression.

## Conclusions

In adults with repaired TOF and preserved LVEF, multimodal imaging identifies an early cardiac remodeling phenotype characterized by atrial remodeling, diastolic dysfunction, and RV systolic dysfunction that is associated with reduced exercise capacity despite the absence of overt myocardial fibrosis. These findings suggest that integration of echocardiographic and CMR markers may improve early detection of physiologic impairment and inform future strategies for risk stratification and therapeutic intervention. These findings support integration of physiologic assessment with imaging-based markers of cardiac remodeling rather than reliance on structural abnormalities alone. Prospective studies are needed to determine whether this approach improves risk stratification or identifies patients who may benefit from earlier intervention.

### Study Limitations

This study has several limitations. Its retrospective design introduces potential selection bias, and the temporal relationship between CPET, TTE, and CMR was not uniform. Assessment of diastolic function in rTOF is inherently challenging due to altered ventricular geometry, conduction abnormalities, and prior surgical interventions, which may affect the interpretation of standard TTE parameters. The composite diastolic dysfunction score, while based on guideline-recommended thresholds, has not been specifically validated in rTOF populations. LGE identifies focal replacement fibrosis but may not detect diffuse interstitial fibrosis; T1 mapping and extracellular volume quantification were not available for systematic assessment in this cohort. CMR was available only in a subset of the cohort, introducing the potential for selection bias in analyses of RV function, ventricular volumes, pulmonary regurgitation, and LGE. Finally, the retrospective cohort design precludes causal inference.

## Clinical Perspectives

### Clinical Implications

These findings have important clinical implications. First, they support consideration of incorporating diastolic parameters and atrial remodeling into the routine assessment of adults with rTOF, as these markers may identify patients with reduced exercise capacity despite preserved LVEF. Second, they suggest that integrating CPET with multimodal imaging may improve identification of patients with early cardiac remodeling before overt myocardial fibrosis becomes detectable. Finally, these findings provide a rationale for investigating whether interventions targeting early diastolic dysfunction and adverse remodeling can preserve functional capacity.

More broadly, these results highlight the value of integrating physiologic assessment with imaging-based markers of cardiac remodeling rather than relying solely on structural abnormalities.

### Translational outlook

Prospective longitudinal studies are needed to determine whether imaging-defined cardiac remodeling predicts progression to overt myocardial fibrosis, ventricular dysfunction, or adverse clinical outcomes in adults with rTOF. Future investigations should evaluate whether integrating CPET with multimodal imaging improves risk stratification and whether interventions targeting early diastolic dysfunction and atrial remodeling can preserve exercise capacity and modify long-term disease progression.

## Data Availability

All data produced in the present work are contained in the manuscript.

## Acknowledgements

The authors thank the Adult Congenital Heart Association for its support of this work through a research grant.

---

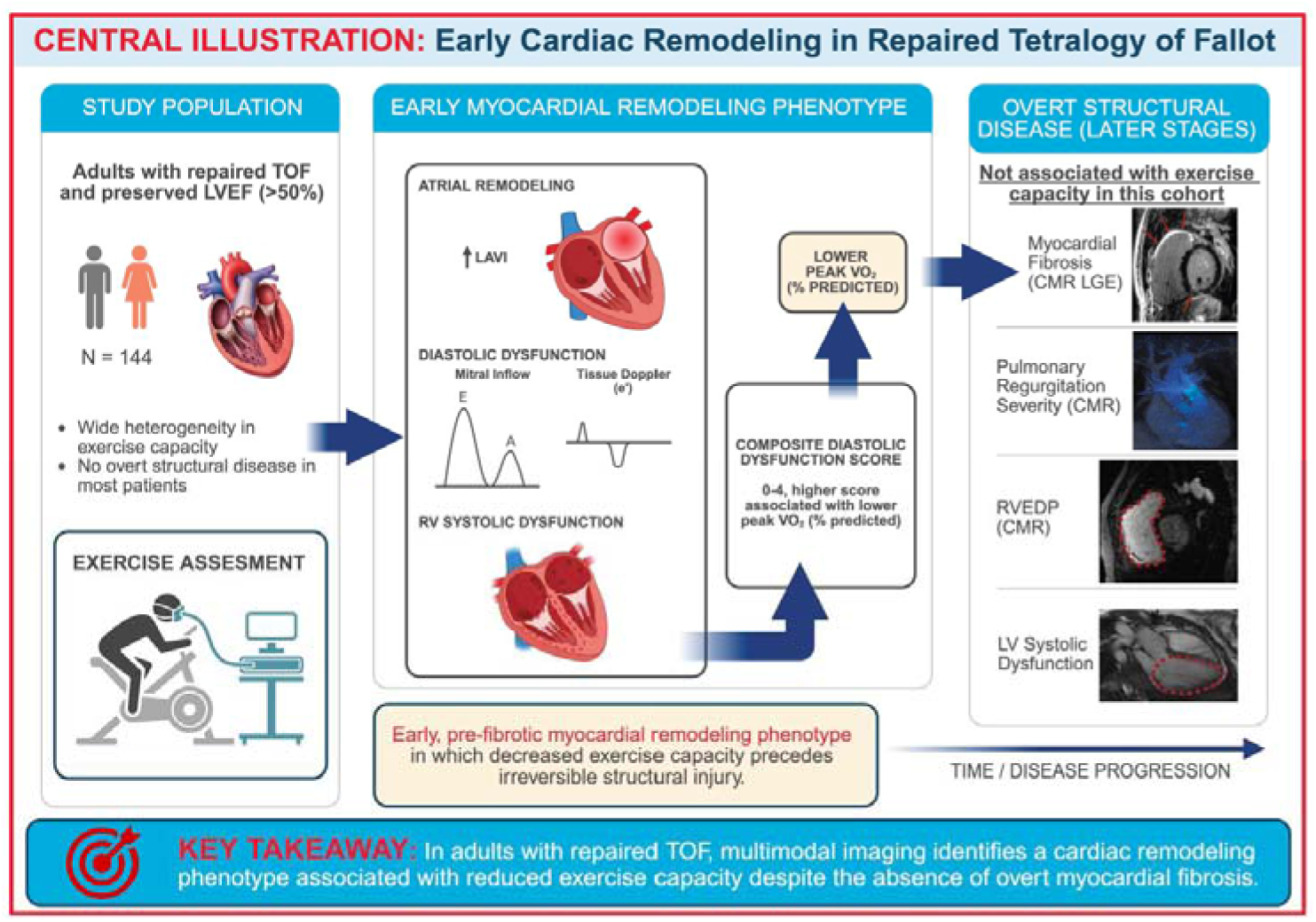
Central Illustration. Cardiac Remodeling and Exercise Capacity in Repaired Tetralogy of Fallot. Adults with rTOF and preserved LVEF demonstrate substantial heterogeneity in exercise capacity despite the absence of overt structural myocardial abnormalities in most patients. Multimodal imaging identified a cardiac remodeling phenotype characterized by atrial remodeling, diastolic dysfunction, and right ventricular systolic dysfunction, each of which was associated with reduced exercise capacity. A higher composite diastolic dysfunction score demonstrated a graded association with lower exercise capacity. In contrast, myocardial fibrosis, pulmonary regurgitation severity, RVEDVi, and left ventricular systolic dysfunction were not associated with exercise capacity in this cohort. CMR = cardiac magnetic resonance; E/e′ = ratio of early transmitral inflow velocity to early diastolic mitral annular tissue velocity; LAVI = left atrial volume index; LGE = late gadolinium enhancement; LVEF = left ventricular ejection fraction; ppVO2 = percent-predicted peak oxygen uptake; rTOF = repaired tetralogy of Fallot; RV = right ventricle; RVEDVi = right ventricular end-diastolic volume index.

## Abbreviations

CMR: cardiac magnetic resonance
CPET: cardiopulmonary exercise testing
E/e′: ratio of early transmitral flow velocity to early diastolic mitral annular velocity
LAVI: left atrial volume index
LGE: late gadolinium enhancement
LVEF: left ventricular ejection fraction
ppVO2: percent-predicted peak oxygen consumption
PR: pulmonary regurgitation
rTOF: repaired tetralogy of Fallot
RV: right ventricle/right ventricular
RVEDVi: right ventricular end-diastolic volume index
RVEF: right ventricular ejection fraction
TR Vmax: peak tricuspid regurgitation velocity

